# How local interactions impact the dynamics of an epidemic

**DOI:** 10.1101/2020.11.24.20237651

**Authors:** Lydia Wren, Alex Best

## Abstract

Susceptible-Infected-Recovered (SIR) models have long formed the basis for exploring epidemiological dynamics in a range of contexts, including infectious disease spread in human populations. Classic SIR models take a mean-field assumption, such that a susceptible individual has an equal chance of catching the disease from any infected individual in the population. In reality, spatial and social structure will drive most instances of disease transmission. Here we explore the impacts of including spatial structure in a simple SIR model. We combine an approximate mathematical model (using a pair approximation) and stochastic simulations to consider the impact of increasingly local interactions on the epidemic. Our key development is to allow not just extremes of ‘local’ (neighbour-to-neighbour) or ‘global’ (random) transmission, but all points in between. We find that even medium degrees of local interactions produce epidemics highly similar to those with entirely global interactions, and only once interactions are predominantly local do epidemics become substantially lower and later. We also show how intervention strategies to impose local interactions on a population must be introduced early if significant impacts are to be seen.

## 1. Introduction

The classic Susceptible-Infected-Recovered (SIR) model has long been used to model the spread of infectious disease in human, animal and plant populations (Kermack and McKendrick, 1927; Anderson and May, 1979). More recently, in its SEIR form (with an additional ‘exposed’ compartment) it has formed a central pillar of much of the modelling of the Covid-19 pandemic (Ferguson et al, 2020; Kucharski et al, 2020; Firth et al, 2020). In its standard form, the SIR model has a mean-field assumption, such that individuals in the population have purely random, ‘global’ interactions (Boots and Sasaki, 2000) and there is no spatial structure. In reality, individuals in a population are more likely to contract disease from infected individuals who are closer to them, both physically and socially. Incorporating this spatial structure into mathematical models is extremely challenging. In some cases, large datasets of known contact networks have been used to replicate epidemics to excellent effect (Ferguson et al, 2020; Firth et al, 2020). While such models have a high degree of realism and thus predictive power, they cannot be readily modelled by a simple set of equations and require significant computational exploration to capture possible outcomes and feedbacks.

One common approach to incorporating a degree of regular spatial structure, and particularly ‘local’ near-neighbour interactions, into infectious disease models is to use a lattice-based probabilistic cellular automata (Sato et al, 1994; Rand et al, 1995). These stochastic individual-based models have also been combined with an analytic pair-approximation method (Matsuda et al, 1992; Sato et al, 1994), where the full spatial dynamics are approximated by a set of ordinary differential equations based on the classic SIR model. Such models have been applied to infectious disease systems both with (Keeling et al, 1997; Webb et al, 2007a,b; Best et al, 2012) and without (Keeling, 1999; Sharkey, 2008) demography. These studies have found that local interactions reduce the value of *R*_0_, slowing or even preventing an epidemic that would occur when interactions are global (Keeling, 1999). These approaches largely insist on a strict degree of spatial structure, where infection and/or host reproduction can only be through near-neighbour interactions. While this is useful for comparison with the mean-field case, interactions are unlikely to be entirely local or global in reality, and we may be missing important features of systems where the interaction structure lies between these two extremes.

The ability to move between local, near-neighbour interactions and global, mean-field interactions has been considered in a few spatial models of infectious disease, primarily in evolutionary (Boots and Sasaki, 1999, 2000; Kamo et al, 2007; Best et al, 2011; Débarre et al, 2012) and ecological (Ellner, 2001; Webb et al, 2007a) contexts. This multiscale method is commonly achieved by allowing a proportion of transmission and/or reproduction to occur locally and the rest globally. We might interpret this, for example in a human population, as an individual mostly interacting within their household or community (local interactions), but also travelling some distance for work, holidays or visiting friends or family (global interactions). These studies have shown that there is increased potential for ecological cycles and disease-driven extinction as interactions become predominantly local (Webb et al, 2007a). They have also shown that evolutionary selection is generally towards lower levels of infection in both host and parasite as interactions become more local (Boots and Sasaki, 1999; Best et al, 2011), but not necessarily monotonically (Kamo et al, 2007). Most recently, this multiscale method has been applied to a human epidemiology model with equal births and deaths (Maltz and Fabricius, 2016), showing that pronounced (but damped) oscillations in infection may result after a sudden shift to local interactions. However, this simple mechanism to investigate the impacts of varying the ‘degree’ of spatial structure (i.e. the relative proportion of local to global interactions) has yet to be applied to human epidemic models over short timescales such that demography does not impact the dynamics, as would be the case in the early stages of an emerging infectious disease.

Given it is known that local spatial structure can rapidly form during epidemics on a network (Keeling, 1999), we would expect local interactions to impact the dynamics of short timescale epidemics, but the impact of introducing different degrees of local interactions in such a model remains unexplored. In this study our aim is to determine how gradually increasing the proportion of local interactions from 0 (completely global) to 1 (completely local) changes the nature of the epidemic. This will allow us to understand how significant movement restrictions - through increasingly local interactions - might need to be to restrict an epidemic. We will also aim to understand the amount of variation that can occur for fixed parameter sets due to the stochastic nature of epidemics, and the extent to which a specific individual-based approximation may be used as a reliable ‘average’ of the stochastic implementations (Sharkey, 2008).

## 2. Model

### 2.1. Mean-field model

The underlying dynamics of the model are based on the classic Susceptible-Infected-Recovered (SIR) epidemiological framework (Kermack and McKendrick, 1927), with no demographic processes (births/deaths). Demographic processes are neglected since we are interested in epidemics on a short time-scale (*<* 12 months) during which we would expect demographics to remain roughly constant. We first consider the model under a mean-field assumption with no local interactions. All individuals in the population are either susceptible (*S*), infected (*I*) or recovered (*R*). The total population size *N* = *S* + *I* + *R* is constant (assume *N* = 1 for consistency with what follows), meaning we only need to track the dynamcis of *S* and *I* densities, given by the following ordinary differential equations,

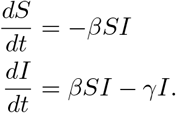

Transmission is assumed to be density-dependent with coefficient *β*, while recovery occurs at rate *γ* and immunity is assumed to be permanent.

### 2.2. Pair-approximation model

To account for spatial structure and local transmission, we use a pair-approximation (PA) model (Matsuda et al, 1992). Assume individuals live on a square lattice, where each site is always occupied by one susceptible, infected or recovered individual. We define the probability that a site is occupied by a susceptible individual as *P*_*S*_, an infected individual as *P*_*I*_ and a recovered individual as *P*_*R*_. The dynamics of these ‘singlet’ densities mirror those of the mean-field model above, with the following ordinary differential equations,

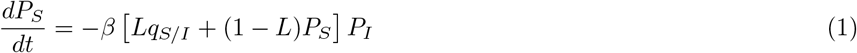

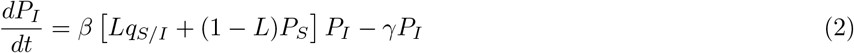

with *P*_*R*_ = 1 *− P*_*S*_ *− P*_*I*_. Here we have introduced our key parameter, *L*, which determines the proportion of transmsision that occurs ‘locally’ between neighbouring individuals, with the remainder of transmission (1 *− L*) occurring ‘globally’ between random individuals on the lattice. This corrersponds to individuals’ interactions being predomiantly local (with their near neighbours) or global (randomly across the population). The conditional probability, called the ‘environs density’, that an infected individual has a neighbour that is susceptible is denoted *q*_*S/I*_ = *P*_*SI*_*/P*_*I*_. Therefore there are two routes to transmission:

- global: (1 − *L*) *β P*_*S*_*P*_*I*_
- local: *Lβ*_*qS/I*_ *P*_*I*_

This system of equations is not closed, since to calculate the conditional probability, *q*_*S/I*_, we need to know the ‘pair’ density, *P*_*SI*_, e.g. the probability that a randomly chosen pair of neighbouring sites are a susceptible and an infected. By considering all possible pair transitions, the dynamics of these pair densities can be expressed as an additional set of ordinary differential equations,

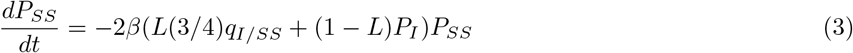

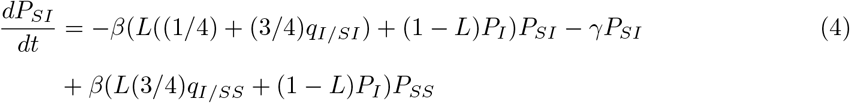

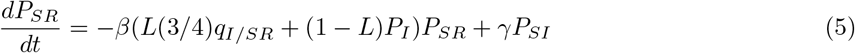

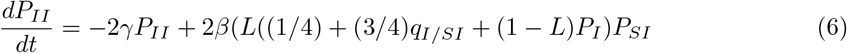

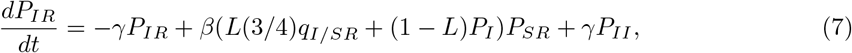

and *P*_*RR*_ = 1 *− P*_*SS*_ *− P*_*II*_ *−* 2*P*_*SI*_ *−* 2*P*_*SR*_ *−* 2*P*_*IR*_. These equations are similar to those of Matsuda et al (1992); Webb et al (2007b); Maltz and Fabricius (2016). This includes further conditional probabilities, for example *q*_*I/SI*_ which is the probability that given we choose an *SI* pair of sites, there is a further neighbour that is an *I* site. Again, this system of equations is not closed as we have these conditional probabilities that depend on ‘triplets’ (e.g. *q*_*I/SI*_ = *P*_*SII*_*/P*_*SI*_). One can appreciate that this pattern will continue and that the equations will never form a closed system. We thus apply a ‘pair approximation’ (Matsuda et al, 1992) where we assume that, for example, *q*_*I/SI*_ = *q*_*I/S*_, allowing us to close the system.

### 2.3. Basic reproductive ratio

The basic reproducive ratio, *R*_0_, is the well-known quantity that measures the average number of secondary infections caused by an infected individual in an otherwise disease-free population (Anderson and May, 1981). Considering the early dynamics of infected individuals (equation (2)), for the mean-field, global case where *L* = 0 (i.e. no local interactions), this is simply given by *R*_0_ = *β/γ*. When interactions are fully local with *L* = 1, we have *R*_0,*l*_ = *βq*_*S/I*_*/γ*. In the limit where the population is indeed entirely disease-free, the conditional density *q*_*S/I*_ = *P*_*S*_ = 1, and the two basic reproductive ratios will be equal.

However, in the early stages of an epidemic the conditional density *q*_*S/I*_ (the probability that an infected individual has a susceptible neighbour) rapidly shrinks as the contact network is formed, since infected hosts will be forming local clusters, meaning it quickly becomes that *R*_0,*l*_ *< R*_0_. This reduction in susceptible contacts and resulting reduction in reproductive ratio naturally leads to a slower epidemic (Matsuda et al, 1992; Keeling, 1999). Given the total reproductive ratio will be,

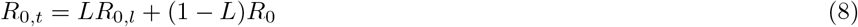

it is clear that the initial growth rate of an epidemic will be slower the greater the degree of local interactions.

### 2.4. Stochastic simulations

Alongside these mathematical models we additionally conduct stochastic individual-based simulations using a probabilistic cellular automata. Similarly to the model described above, a lattice of sites is established, now of fixed size (25×25), where each site is again occupied by one individual. A Gillespie algorithm (Gillespie, 1977) is implemented, where waiting times between events (either recovery (*I → R*) or local or global transmission (*S → I*)) are drawn from an exponential distribution with mean given by the sum of the total rates (e.g. *γI* + *βSI*). At each step, exactly one of these events occurs, with probabilities proportional to their rates, and a suitable host is chosen randomly from the lattice for it to occur to (e.g. recovery requires an infected host to be selected). After an event occurs, the lattice is updated and a new waiting time calculated for the next event. This approach is fully spatially explicit, unlike the approximation present in the mathematical methods above. It also now has a discrete number of individuals (625) as opposed to the continuous probabilties in the PA model. Code is available on https://github.com/abestshef/latticeSIR.

## 3. Results

### 3.1. Epidemic curves

We begin with a visual examination of the epidemic curves predicted by the pair approximation and stochastic simulations for different values of *L* (0.1 and 0.9) and different mean-field basic reproductive ratios, *R*_0_ (2, 5 and 10). Recent work has highlighted the problems of combining mutliple stochastic individual-based models into simple static statistics of means and variances (Juul et al, 2020). We follow the methods of Juul et al (2020) by finding the ‘most central’ 50% of 100 simulated curves to present here (see appendix for details). Below we provide further detail by examining three key descriptive statistics of the epidemic.

Focussing on the effect of increasing the proportion of local interactions, from figure 1 it is clear viusally that the higher value of *L* produces a lower and later peak of infection. Restricting global interactions may therefore, in itself (without further reductions to transmission probability), slow down and limit the spread of an epidemic. Increasing *R*_0_ not only moves the epidemics earlier and higher, but also reduces the effect of local interactions. Comparing the plots, we can see that control mechanisms that both shift interactions from predominatly global to predominatly local *and* reduce *R*_0_ (for example, through both movement restrictions and other non-pharmaceutical interventions) are predicted to have a significant effect on reducing the epidemic.

**Figure 1.**
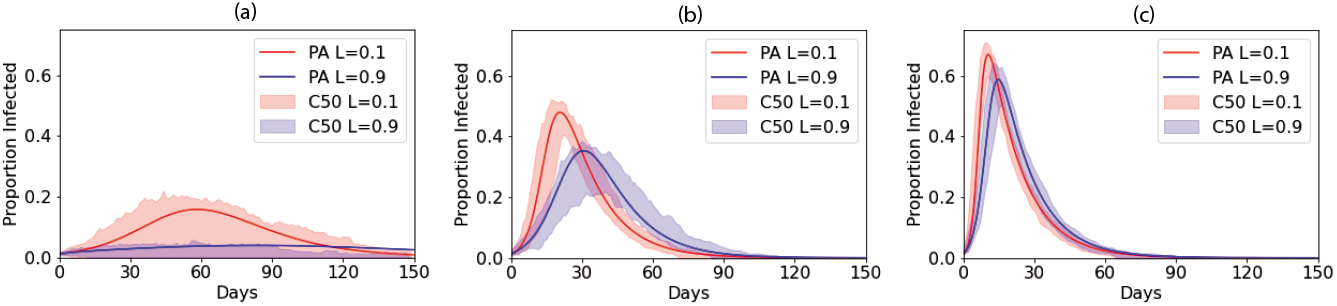
Epidemic curves from pair approximations and the ‘most central’ 50% of 100 stochastic simulations for different values of *L. γ* = 1*/*14. (a) *R*_0_ = 2, (b) *R*_0_ = 5, (c) *R*_0_ = 10. Red curves for *L* = 0.1 and blue curves for *L* = 0.9. The solid line shows the PA dynamics and the shading the bounds of the central 50% (C50) of runs of the stochastic model.

We can also compare the fit of the pair-approximation to the stochastic models. As we might expect, when *L* is small the pair approximation appears to present a reasonable ‘average’ of the stochastic model runs. As *L* becomes larger we find that, while the pair approximation often sits within the most central runs, for larger *R*_0_ at least, it tends to predict that the epidemic peak is rather earlier and higher than seen in most of the fully spatially-explicit simulations. The discrepancy between the pair approximation and stochastic simulations is most pronounced at low values of *R*_0_. In particular, in this case a number of the stochastic simulations produce ‘failed’ epidemics, as evidenced by the lower bound of the 50% central curves running close to 0.

### 3.2. Descriptive statistics

We now explore the behaviour as we vary local interactions across the full range of *L* from 0 (fully global) to 1 (fully local). Three descriptive statistics were evaluated (percentage of the population infected by day 300, percentage of the population infected at the peak and the day of the peak). These not only provide a useful summary of the epidemic curve (*‘How high was it? How long did it last?’*) but are also potential targets in setting public health policy (i.e. we may not wish the peak to pass a certain threshold). The stochastic model was run 100 times and the results plotted using boxplots, showing the median, inter-quartile range (IQR; 25%-75%), maxima/minima (or 1.5*×*IQR if smaller) and outliers. We consider how similar results are to the *L* = 0 case by noting where the IQRs do and do not overlap. We further conduct pairwise Z-tests to compare the means of the simulation results, presenting the resulting p-values in the appendix. We set significance thresholds of 5% and 1%, with a Bonferroni correction for multiple tests (n=55 pairwise tests).

Two clear trends emerge from all of the results. Firstly, there is an accelerating impact of local interactions, with little effect seen as *L* is first increased from 0, but the impact growing as *L* moves towards 1. This is highlighted by the colours indicating which cases have overlapping IQRs with the *L* = 0 case (the bounds of which are shown using horizontal dashed lines). For all three measures there are overlapping IQRs at least up to *L* = 0.4, and often higher, indicating that the output for these cases is similar to the *L* = 0 case. As *L* reaches higher values there are then rapid moves away from the *L* = 0 case towards smaller epidemics (with a more complex impact on the day of the peak; see below). Pairwise Z-tests (see appendix) confirm that *L* = 0.1 and *L* = 0.2 are never significantly different to the *L* = 0 mean for peak infections (p*>*0.01/55), with significant differences for all *L* ≥ 0.6 for both peak and total infected (p*<*0.01/55). In contrast, in only one measure (peak infected, *R*_0_ = 2) is the *L* = 1 mean not significantly different from all other cases. Secondly, the impact of local interactions is reduced for higher *R*_0_. For every statistic, the number of overlapping IQRs increases for *R*_0_ = 10 compared to *R*_0_ = 2.

Focussing on the specific values, when *R*_0_ = 2 increasing the proportion of local interactions from *L* = 0 to *L* = 0.5 reduces the median peak from 17% to 13%, but increasing further to *L* = 1 reduces it to just 2% (figure 2a). Similarly, the median total infected is reduced when changing *L* = 0 to *L* = 0.5 from 80% to 70%, but at *L* = 1 it is reduced to only 6% infected (figure 2g). Similar patterns for the peak can be seen from higher *R*_0_ values also, with *R*_0_ = 5 seeing median peaks of 49% for *L* = 0 reduced first to 47% for *L* = 0.5 then to 20% for *L* = 1 (figure 2b), while for *R*_0_ = 10 the median peak of 68% at *L* = 0 is reduced just to 67% for *L* = 0.5 but 52% for *L* = 1 (figure 2c). The patterns for total infected, however, are less pronounced at higher *R*_0_.

**Figure 2.**
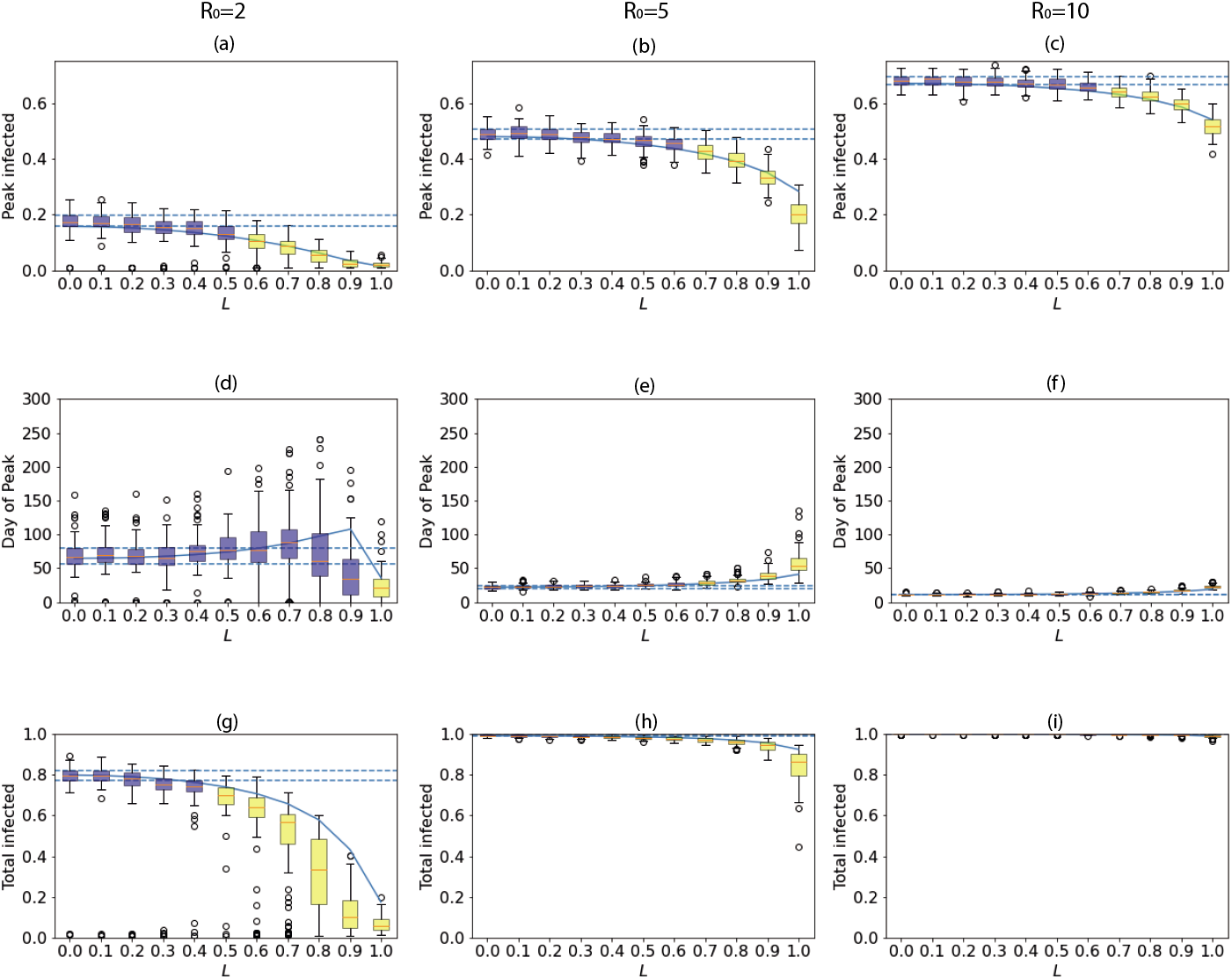
Descriptive statistics of PA and stochastic simulations for (left) *R*_0_ = 2, (middle) *R*_0_ = 5 and (right) *R*_0_ = 10. (a)-(c) Proportion infected at peak, (d)-(f) Day of peak, (g)-(i) Proportion of population infected by day 300. Results from 100 simulations are presented as boxplots highlighting the median (orange lines), inter-quartile ranges (IQR; boxes), maxima/minima (or 1.5 *×* IQR if smaller; whiskers) and outliers. Dashed horizontal lines mark the bounds of the IQR for *L* = 0. Blue boxes have overlapping IQRs with *L* = 0 while yellow boxes do not. The solid line marks the PA.

Figure 2(d)-(f) shows that the number of days until the peak increases with *L*, again accelerating as *L* increases. There is an exception to this when *R*_0_ = 2 as *L* approaches 1. Here, the peak moves significantly earlier because the infection fails to spread through the population meaning the peak of the epidemic is both very early and very low, as confirmed in figure 2a. Obviously, the larger *R*_0_ is, the faster the disease will be able to spread through the population and therefore the faster it will die out.

In general, the pair approximation appears to be a good ‘average’ of the results from the stochastic mode since it is always within the maximum/minimum bounds and regularly within the IQRs. The ‘fit’ appears to be least good as *L* approaches 1, as would be expected. The pair approximation is less accurate for *R*_0_ = 2 than for higher values of *R*_0_, and this is likely due to the large proportion of infections which fail to become established in the stochastic model when the disease spreads slowly, resulting in lower means and IQRs, as described in the online appendix. The pair approximation also fails to account for variation evident for some of the stochastic simulations. For example, for *R*_0_ = 2 and *L* = 0.8 the total proportion infected can be anything from almost 0% to more than 60%, whereas the pair approximation, as a deterministic approximation, provides just a single value.

### 3.3. Using local interactions as a control mechanism

We now explore how enforcing movement restrictions, resulting in more localised interactions, might impact the spread of an epidemic. We assume that initially a population has predominantly global interactions (*L* = 0.1). We then assume that when a threshold of percentage infected (here, 5%) is reached, interactions immediately switch to being predominantly local (*L* = 0.9) and remain so until the infected percentage returns below the threshold. Figure 3 shows that compared to the case where interactions remain predominantly global throughout (red), if movement restrictions are imposed (blue) the peak of the epidemic is reduced, but less substantially than if interactions had always been predmoniantly local, particularly for the lower *R*_0_ (see figure 1 and table 1). Interestingly, we see a very slight second wave emerging for lower *R*_0_ once restrictions are lifted since the herd-immunity threshold has not been reached, suggesting further and/or longer restrictions may need to be imposed. We further investigate by varying the threshold at which restrictions are imposed and the value of *L* moved to under the restrictions (figure 4). It is clear a lower threshold improves the ability to control the epidemic. However, while reducing the threshold has an almost linear effect on the peak, a very low threshold is needed to impact the total infected with higher threshold making only small changes (figure 4 and table 1).

**Table 1:** The median peak and total infected for different threshold proportions of infection when the proportion of local interactions is moved from *L* = 0.1 to *L* = 0.9 with *R*_0_ = 2.

| Threshold | Median peak | Median total |
| --- | --- | --- |
| None | 17% | 80% |
| 10% | 13% | 73% |
| 5% | 9% | 66% |
| 1% | 4% | 36% |

**Figure 3.**
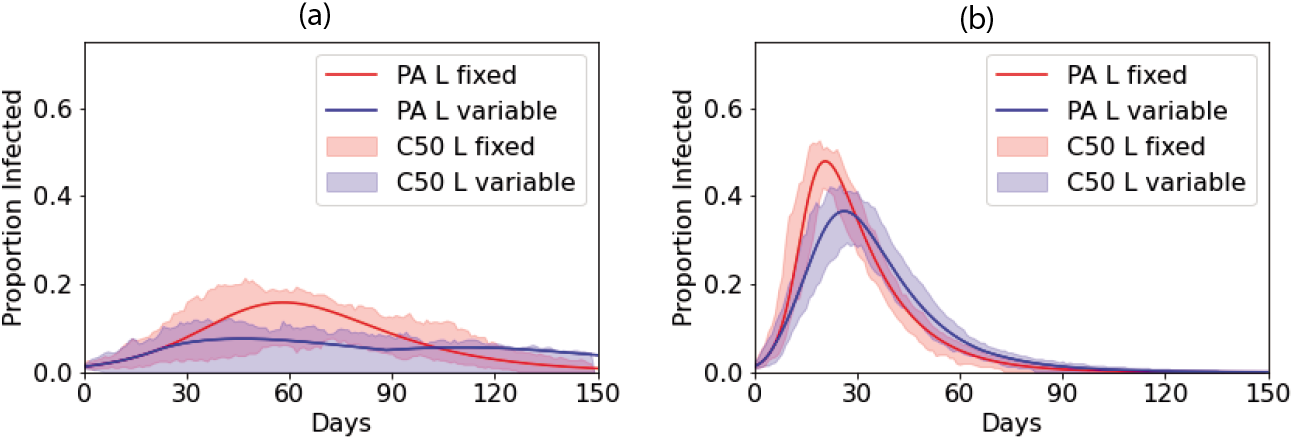
Central curves and PA with (a) *R*_0_ = 2 and (b) *R*_0_ = 5. Red curves, *L* = 0.1 throughout. Blue curves: *L* = 0.1 until *P*_*I*_ *>* 0.05, then *L* = 0.9 while *P*_*I*_ *>* 0.05, dropping back to *L* = 0.1 thereafter.

**Figure 4.**
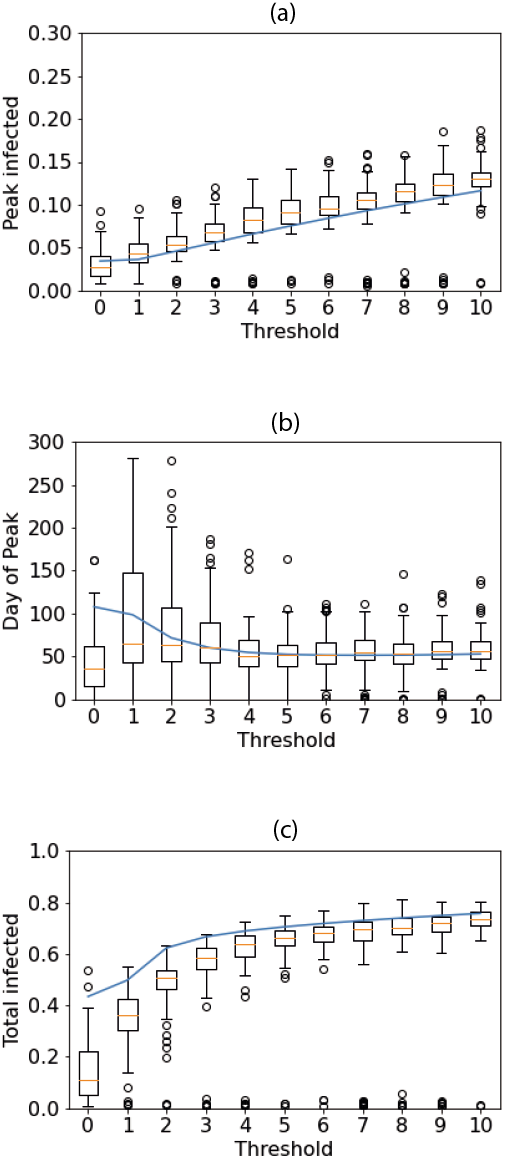
Impact of changing the threshold at which interactions switch from *L* = 0.1 to *L* = 0.9 with *R*_0_ = 2. Boxplots show results from stochastic simulations and solid line the PA.

This relative lack of impact of later interventions is because of the speed with which the lattice becomes correlated in the early stages of an epidemic. The correlation between *S* and *I* sites on the lattice - effectively how likely it is to find *S* sites next to *I* sites - is given by,

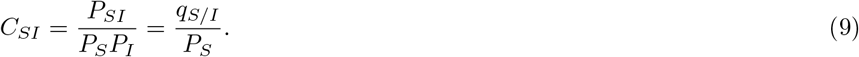

At the start of an epidemic with predominantly global interactions, the lattice is uncorrelated since infection spreads randomly across the lattice. As such, an infected host is likely to remain surrounded by susceptible individuals and therefore *C*_*SI*_ = 1. If some interactions are local, then during the early stages the correlation rapidly approaches a quasi-equilibrium as the contact network forms and local patches of infection develop (Keeling, 1999). We show in the appendix that this can be approximated as,

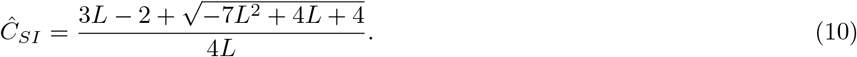

Figure 5 shows that increasing *L* leads to much stronger early-time *S*-*I* correlation, as local clusters of infection form due to spatially-localised contact networks, meaning that *I* individuals are much more likely to be located near other *I* individuals. If an epidemic begins in a population with predominatly local interactions, the lattice quickly becomes correlated with these local clusters of infection, *q*_*S/I*_ falls and the infection slows itself down due to a lack of locally available susceptible individuals. In contrast, if an epidemic has established with predominatly global interactions, the network is already highly uncorrelated before the movement restrictions are imposed and there is already infection spread widely across the lattice, meaning most *I* individuals have many *S* neighbours. The late implementation of local interactions therefore cannot cause as high correlation of the lattice, and a large number of local epidemics can still occur.

**Figure 5.**
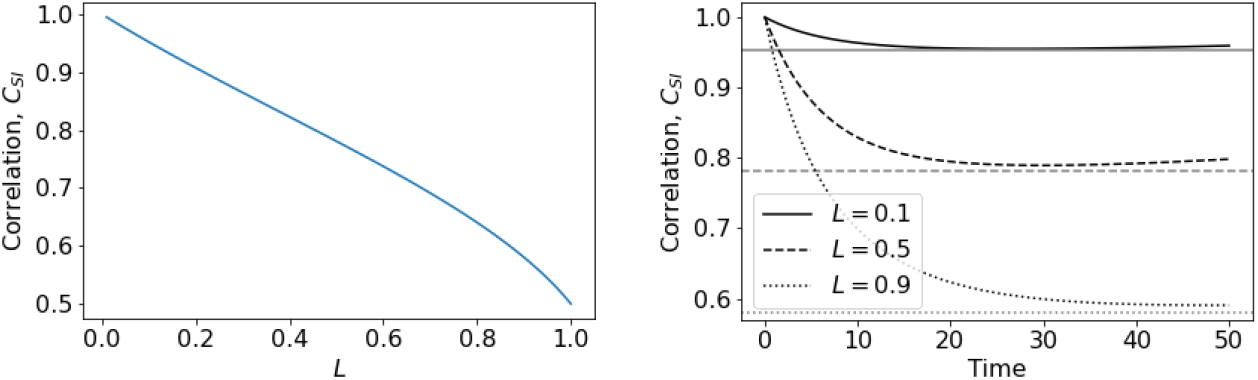
Correlation coefficient, *C*_*SI*_, from pair approximation for different values of *L*. Left: Predicted quasi-equilibrium, *Ĉ*_*SI*_ from equation (10). Right: Early-time correlation dynamics *C*_*SI*_ from full pair-approximation model. The grey horizontal lines mark the predicted quasi-equilbrium from equation (10).

## 4. Discussion

In this study we have used a pair approximation alongside stochastic simulations to investigate the impact of local interactions on an epidemic. The novelty of our model is to explore how epidemics over short timescales can be restricted by different degrees of local interactions, not just at the extremes of purely global or purely local infections. Our results show that epidemics where interactions are predominantly local will result in fewer infections than those where interactions are predominantly global. Moving from fully global to fully local interactions could reduce the median total infected from 80% to just 6% in one case. This is in line with previous studies that looked only at the extreme cases (Keeling, 1999). Importantly, however, we have investigated the transition between these extremes, finding that the trends as we move from purely global to purely local interactions are not linear. Instead, our results consistenly show initially flat responses in different infection statistics as *L* is increased, with rapid changes as *L* approaches 1 (figure 2). This suggests that the course of an epidemic in a population with relatively high proportions of local interactions (even 50:50) will be roughly the same as an epidemic in a population with purely global interactions. Even at relatively low proportions of global interactions, enough long-range infections can occur in the early stages of an epidemic to seed large numbers of local epidemics, allowing the infection to spread throughout the population. For example, if *R*_0_ = 2 and *L* = 0.5, on average an infected individual passes the disease to one local and one global contact, allowing the disease to become established across the lattice and to then form a series of outbreaks. It is only as *L* becomes close to 1 and almost all interactions are local that the likelihood that an infected individual transmits the disease globally is small enough to have a significant impact. Interestingly, in the similar model by Maltz and Fabricius (2016) that includes simple demographics (and thus yields an endemic equilbrium), the infected equilibrium is initially fairly static as interactions become more local before rapidly falling as local interactions become more dominant, suggesting this non-linear trend is robust in simple epidemic models.

Our results have important implications for attempting to limit an epidemic through restricting movement. In particular, such restrictions must be considerable, with almost all global interactions removed if significant effects are to be seen. It is important to note that in our model restricting movement does not lead to lowered per-individual contacts, as might be assumed under simple non-pharmaceuitcal interventions (for example social distancing, hand hygiene, wearing masks). We found that restrictions that both make interactions more local and infectious contacts less frequent (through lowered *R*_0_) can substantially reduce the impact of an epidemic. Moreover, we found that if the population starts from a position of having predominantly global interactions, movement restrictions must be imposed very early on in the course of an epidemic or they will have minimal effect (figure 4). This is due to the fact that, if a disease has already begun to spread randomly through a population with global contacts, when restrictions are put in place there will already be large numbers of local outbreaks forming across the lattice. If an infection has a particularly high *R*_0_, and therefore rapid growth, it may be that infection is already too widespread for movement restrictions to have any effect by the time measures are implemented. In this study we assumed a simple switch such that interactions returned to the default after the infected proportion fell back below the threshold. More realistic approaches might be to gradually ease restrictions or enact further restrictions in cases where a ‘2nd wave’ emerges. In the similar study by Maltz and Fabricius (2016), they found a simple switch to a different proportion of local interactions led to pronounced (damped) oscillations and significant periodic outbreaks as the system was effectively moved such that it was no longer at its steady state. Further investigation in to the use of movement restrictions as a control mechanism is needed to explore the best strategies.

Combining mathematical analysis, using the pair approximation (Matsuda et al, 1992; Sato et al, 1994), and stochastic simulations has allowed us to explore the dynamics of our model in more depth. Interestingly we found that the deterministic results from the pair approximation model provide a good ‘average’ of the dynamics from fully-spatial stochastic simulations. The weakest ‘fits’ were for our lowest values of *R*_0_, where a proportion of simulations lead to failed epidemics, whereas the analytical model always assumes an outbreak occurs. Given the problems in accurately depicting averages of stochastic simulations (Juul et al, 2020), such analytic approximations may provide a useful guide. However, we did find cases where significant variation was present in the stochastic simulations, with the total infected varying from almost 0 to 60% for certain parameter sets, and the pair approximation is not able to capture such variation. The use of the pair approximation did, however, allow us to approximate the correlation of *S* and *I* sites and therefore determine why late interventions did not succeed in restricting the epidemic.

We have deliberately focussed on the simplest possible epidemic model in this study, with the only two mechanisms being transmission and recovery. This has allowed us to draw clear conclusions and insight in to the behaviour of the model, but it clearly cannot and should not be used as an accurate predictive model for a particular epidemic. In an earlier study, Maltz and Fabricius (2016) considered the same model with simple demographics, finding that the infected equilibrium reduces with more local contacts, while (Webb et al, 2007a) examined the impact of varying local interactions on a fully ecological model, noting the potential for disease-induced extinctions and endemic cycles of disease. Clearly, however, there are many further elements that could be considered to make the model appropriate for specific infections or systems. A standard extension for many disease models is to add an exposed compartment, separating out those that are infected from those that are also infectious (see Keeling and Rohani, 2008). It may also be instructive to consider the dynamics if immunity to infection wanes over time, since the non-spatial model would then yield an endemic equilibrium, unlike our model. If we wish to consider a disease persisting over the long-term, we should not only add demographics but also consider seasonal-forcing (Aron and Schartz, 1984; Schwartz, 1985; Altizer et al, 2006). Finally, more realistic spatial and social networks would be needed for any conclusions around movement/interaction restrictions in specific circumstances to be considered, such as in recent models of Covid-19 in the UK (Ferguson et al, 2020; Kucharski et al, 2020; Firth et al, 2020). As it is, our model suggests that significant movement restrictions may be a useful strategy in tackling an epidemic.

## Data Availability

N/A

## Acknowledgements

Thanks to Ben Ashby and Alison Poulston for helpful discussions when developing the project, and to four anonymous reviewers for their feedback on an earlier version of the manuscript. LW received an Undergraduate Research Internship stipend from The School of Mathematics and Statistics at Sheffield University.

## Availability of data

None.

## Availability of code

Code is available from https://github.com/abestshef/latticeSIR.

## Author contributions

AB conceived of and designed the study. LW and AB developed and analysed the model and wrote the manuscript.

