## Appendix for "How local interactions impact the dynamics of an epidemic" (from another version of this preprint: version 1, 2020-11-28)

### A. Plotting 'most central' simulation runs

Recent work has highlighted that static statistics of epidemic curves can provide a poor guide to actual outcomes (Juul et al., 2020; see main text for reference). We did not use the package developed by these authors but implemented our own code, closely following their methods, for ease of application to our own output. The alogorithm was as follows:

(1) 100 simulated epidemic curves were produced from the stochastic model for a certain parameter set, and the time course discretised to integer units.
(2) Scores of every curve were set to 0.
(3) 20 of the curves were randomly selected.
(4) The envelope of the 20 curves was found (i.e. the maximum and minimum of the 20 curves at every time point).
(5) If any curve fell entirely within the envelope for all time points, its score was increased by 1.
(6) Steps 2-4 were repeated 50 times.
(7) The curves were then ranked by their scores, and the top 50% selected for plotting.

### B. Calculation of early-time correlation quasi-equilibrium

As stated, we have,

$$C_{SI} = \frac{P_{SI}}{P_S P_I}. \tag{B.1}$$

Therefore,

$$\frac{dC_{SI}}{dt} = \frac{1}{P_S P_I}\left(\frac{dP_{SI}}{dt} - C_{SI}\left(P_S\frac{dP_I}{dt} + P_I\frac{dP_S}{dt}\right)\right). \tag{B.2}$$

$$= \beta\left(\left(\left(P_S\frac{q_{I/S}}{4} - q_{S/I} + \frac{3q_{S/S}}{4} - \frac{1}{4}\right)L - P_S\right)C_{SI} + (1-L)q_{S/S}\right). \tag{B.3}$$

Early in the epidemic we can assume that $P_S \approx 1, P_I \approx 0, q_{S/S} \approx 1$ and $q_{I/S} \approx 0$. This then simplifies the dynamics to,

$$\frac{dC_{SI}}{dt} = \beta\left(\left(\left(\frac{3}{2} - C_{SI}\right)L - 1\right)C_{SI} + (1-L)\right), \tag{B.4}$$

the solution of which is as given in the main text.

### C. Additional plots

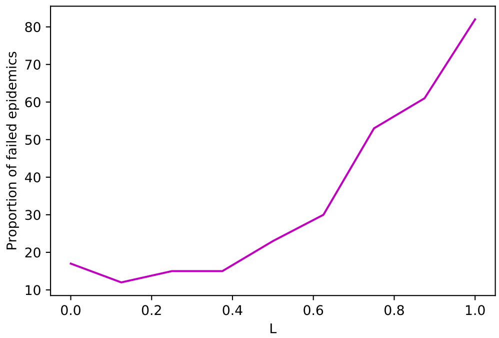


FIGURE 1. Proportion of 'failed' epidemics, defined as the infection dying out before day 100, for different values of $L$ with $R_0 = 2$.

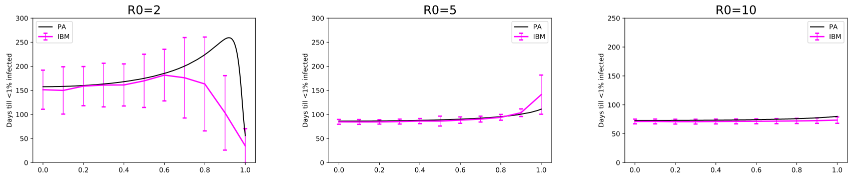


FIGURE 2. Days until less than 1% of population are infected for varying $L$ and three different values of $R_0$.

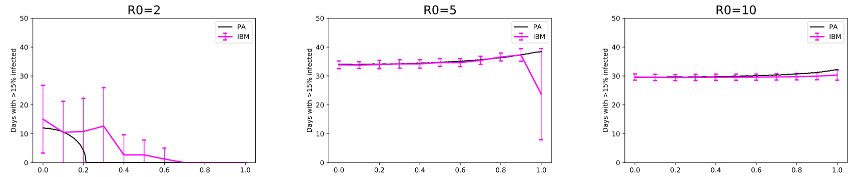


FIGURE 3. Days with more than 15% of population infected for varying $L$ and three different values of $R_0$.
